# Change in Striatal Functional Connectivity Networks Across Two Years Due to Stimulant Exposure in Childhood ADHD: Results from the ABCD Sample

**DOI:** 10.1101/2024.03.18.24304470

**Authors:** Adam Kaminski, Hua Xie, Brylee Hawkins, Chandan J. Vaidya

## Abstract

Widely prescribed for Attention-Deficit/Hyperactivity Disorder (ADHD), stimulants (e.g., methylphenidate) have been studied for their chronic effects on the brain in prospective designs controlling dosage and adherence. While controlled approaches are essential, they do not approximate real-world stimulant exposure contexts where medication interruptions, dosage non-compliance, and polypharmacy are common. Brain changes in real-world conditions are largely unexplored. To fill this gap, we capitalized on the observational design of the Adolescent Brain Cognitive Development (ABCD) study to examine effects of stimulants on large-scale bilateral cortical networks’ resting-state functional connectivity (rs-FC) with 6 striatal regions (left and right caudate, putamen, and nucleus accumbens) across two years in children with ADHD. Bayesian hierarchical regressions revealed associations between stimulant exposure and change in rs-FC of multiple striatal-cortical networks, affiliated with executive and visuo-motor control, which were not driven by general psychotropic medication. Of these connections, three were selective to stimulants versus stimulant naive: reduced rs-FC between caudate and frontoparietal network, and between putamen and frontoparietal and visual networks. Comparison with typically developing children in the ABCD sample revealed stronger rs-FC reduction in stimulant-exposed children for putamen and frontoparietal and visual networks, suggesting a normalizing effect of stimulants. 14% of stimulant-exposed children demonstrated reliable reduction in ADHD symptoms, and were distinguished by stronger rs-FC reduction between right putamen and visual network. Thus, stimulant exposure for a two-year period under real-world conditions modulated striatal-cortical functional networks broadly, had a normalizing effect on a subset of networks, and was associated with potential therapeutic effects involving visual attentional control.

## INTRODUCTION

Prescribed since the 1950s, stimulant medications (e.g., amphetamines, methylphenidate) have well-established efficacy for the acute attenuation of hyperactivity, impulsivity, and inattention, the core symptoms of Attention Deficit Hyperactivity Disorder (ADHD) (1,2). While stimulants are the first-line treatment for ADHD, used by an estimated 3.5% of children in the US (3,4), their long-term effects on the brain are not well understood. Carefully controlled prospective studies investigating stimulant exposure in ADHD have identified modulation in resting-state functional connectivity (rs-FC), measured by the temporal correlation of neural activity in a task-free state, which is associated with chronic stimulant exposure and can partially explain symptom reduction among stimulant-treated children and adolescents (5–7). While controlled prospective studies are essential for teasing apart the relationships between stimulant exposure, functional characteristics of the brain, and heterogeneous symptom trajectories, the study protocols of controlled trials do not exactly match circumstances of real-world community treatment, which are marked by high nonadherence leading to intermittent stimulant exposure (8–10) and multiple psychotropic medication prescriptions (11). Medication adherence, described as taking medication in a way “corresponding to agreed recommendations from a clinician” (12), is particularly poor for adolescents with ADHD, with estimations ranging from 13.2% to 64% (8,13). The effects of stimulant exposure in the context of uncontrolled community-based treatment are largely unknown. While long needed, community datasets that allow examination of real-world neural and symptom changes longitudinally in children being treated for ADHD with stimulants have not been available. Such an opportunity is now at hand with the Adolescent Brain Cognitive Development (ABCD) study (14), currently the largest multisite brain imaging project recruiting children in the US at age 9 years (baseline) who are assessed every two years with functional and structural brain imaging and gold standard clinical assessments.

Rs-FC approaches have revealed the existence of large-scale, distributed, and functionally-specialized networks in the brain (e.g., the default mode network, active during rest; canonical executive networks [e.g., the frontoparietal network], active during tasks) which have emerged as key loci of dysfunction in psychiatric disorders (15,16) and are disrupted in ADHD (17,18). Prospective controlled studies reveal that stimulant-associated modulation of cortical network rs-FC, notably with striatal regions, may explain ADHD symptom change over periods of weeks to years (referred to as chronic). For instance, strengthened rs-FC between the default mode network and putamen after 6 months of stimulant treatment was shown to relate to reduction in ADHD symptoms in childhood (5). In adolescents, reduced rs-FC between canonical executive network regions and the caudate was shown to relate to attentional performance and good response to treatment following an 8-week methylphenidate trial (19). In adults, greater reduction in rs-FC between executive network regions and dorsal anterior cingulate as well as insular cortex was shown to relate to greater reduction in ADHD symptoms after 3 weeks of treatment (7). Similarly, one observational study found that reduction in rs-FC between multiple executive network regions was related to better response to stimulant exposure, following multiple assessments ranging from 7 to 17 years old (20). Together, these findings suggest that chronic stimulant exposure, especially in a controlled context, results in modulation of rs-FC, notably a reduction in the rs-FC of canonical executive networks, and that this modulation may distinguish responders from non-responders (see review 21). While these studies highlight important progress, small sample sizes and methodological heterogeneity obscure firm conclusions (21). More importantly, effects of chronic uncontrolled stimulant exposure matching the conditions of real life, such as possibly inconsistent medication adherence and polypharmacy, remain largely unknown.

ADHD is marked by heterogeneity in long-term symptom trajectories, even for stimulant-treated adolescents, which obscures group-level effects. For example, while it has been reported that treatment with stimulants may not alter long-term trajectories of ADHD symptoms, socioemotional functioning, or motor control in children with ADHD (22), the considerable variability in treatment response may drive the group-level effects to be weaker (23) and highlights the need to parse individual symptom courses. Symptoms for some individuals, as demonstrated by prospective longitudinal studies, even persist into adolescence and adulthood with no demonstrable benefit of stimulant medication on symptom severity or overall functioning (24–26). Because only a subset of children with ADHD will respond to stimulant treatment, identifying the neurobiological features which explain this heterogeneous response to stimulants is an important step for personalizing ADHD therapies. Whether treatment personalization may be feasible also importantly depends upon understanding stimulant-response in the context of uncontrolled naturalistic settings.

Here, we examined striatal-cortical rs-FC change and its association with both stimulant exposure and reliable ADHD symptom improvement across two years of the first ABCD cohort. As one of the largest pediatric community datasets (n=11,878) with neuroimaging data, ABCD offers the best possible opportunity to test stimulant exposure in real-world conditions. We examined the striatum, a direct target of stimulant action by dopamine transport blockade (27), which has consistently shown structural and functional atypicalities in children and adults with ADHD (17,28), and focused on large-scale functional networks as they are known to be sensitive to developmental pathophysiology and stimulant exposure. Since the ABCD study is observational, dosage and duration of medication exposure was not controlled. For rigor we parsed stimulant-specific effects in a three-pronged analytic approach: first, we contrasted participants with ADHD whose parents or guardians reported use of stimulants during the two-year period with those reported to not be using them. To address the expected presence of comorbidity and polypharmacy in the sample (29), we additionally wanted to eliminate the possibility of effects being driven by psychotropic medication generally, without limiting the sample size beyond applying strict exclusion criteria for head motion. We therefore split the stimulant naive group into individuals reported to be taking other psychotropic medications (e.g., antidepressants) and individuals reported to not be taking any psychotropic medication. Using seed-based rs-FC of left and right caudate, putamen, and nucleus accumbens with 10 bilateral large-scale cortical networks, we identified rs-FC changes across two years which predicted stimulant exposure but *did not predict* other psychotropic medication exposure. Second, selecting only connections passing these two criteria, we next identified which of these connections changed between baseline and the 2-year timepoint in the stimulant-exposed group but not in the stimulant-naive group. We also compared rs-FC change of the stimulant-exposed group to children in the ABCD sample who had no psychiatric diagnosis (a typically developing group), in order to determine whether or not these changes suggested normalization of rs-FC. Third, for connections passing this second step, we tested whether rs-FC change predicted reliable symptom improvement. We employed Bayesian multilevel (BML) modeling for all steps in an effort to efficiently address the issue of multiple testing without over-penalizing effect estimates, as this method has been shown to deal with multiplicity in imaging data while controlling for incorrect sign and magnitude error (30).

## MATERIALS AND METHODS

### Participants

Data from 11,878 individuals in the Adolescent Brain Cognitive Development (ABCD) Study (14,31) were obtained from the National Institute of Mental Health Data Archive, ABCD 4.0 data release. From this sample, we limited inclusion to children who met the following criteria: 1) ADHD diagnosis at baseline based on the parent/guardian-reported Kiddie Schedule for Affective Disorders and Schizophrenia (KSADS-5; n=1085 out of 11878; (32)), 2) T-score > 60, a cutoff indicative of risk for clinical symptoms (60–64) or clinical symptoms (>=65), on the DSM-oriented ADHD Problems Scale of the parent/guardian-reported Child Behavior Checklist (CBCL) at baseline (n=734 out of 1085; (33,34)), 3) ABCD Study recommended rs-FC inclusion criteria based on head motion and the quality of FreeSurfer reconstruction (35) at baseline and year two (Y2) timepoints (n=202 out of 734), and 4) had mean framewise displacement (FD) less than 0.5mm for both baseline and Y2 resting-state data, in order to minimize the effect of head motion (n=179 out of 202). These criteria resulted in a sample of 179 individuals (mean age at baseline=9.91 years, at Y2=11.90 years; 69 Female), which is described in Table 1.

**Table 1.**
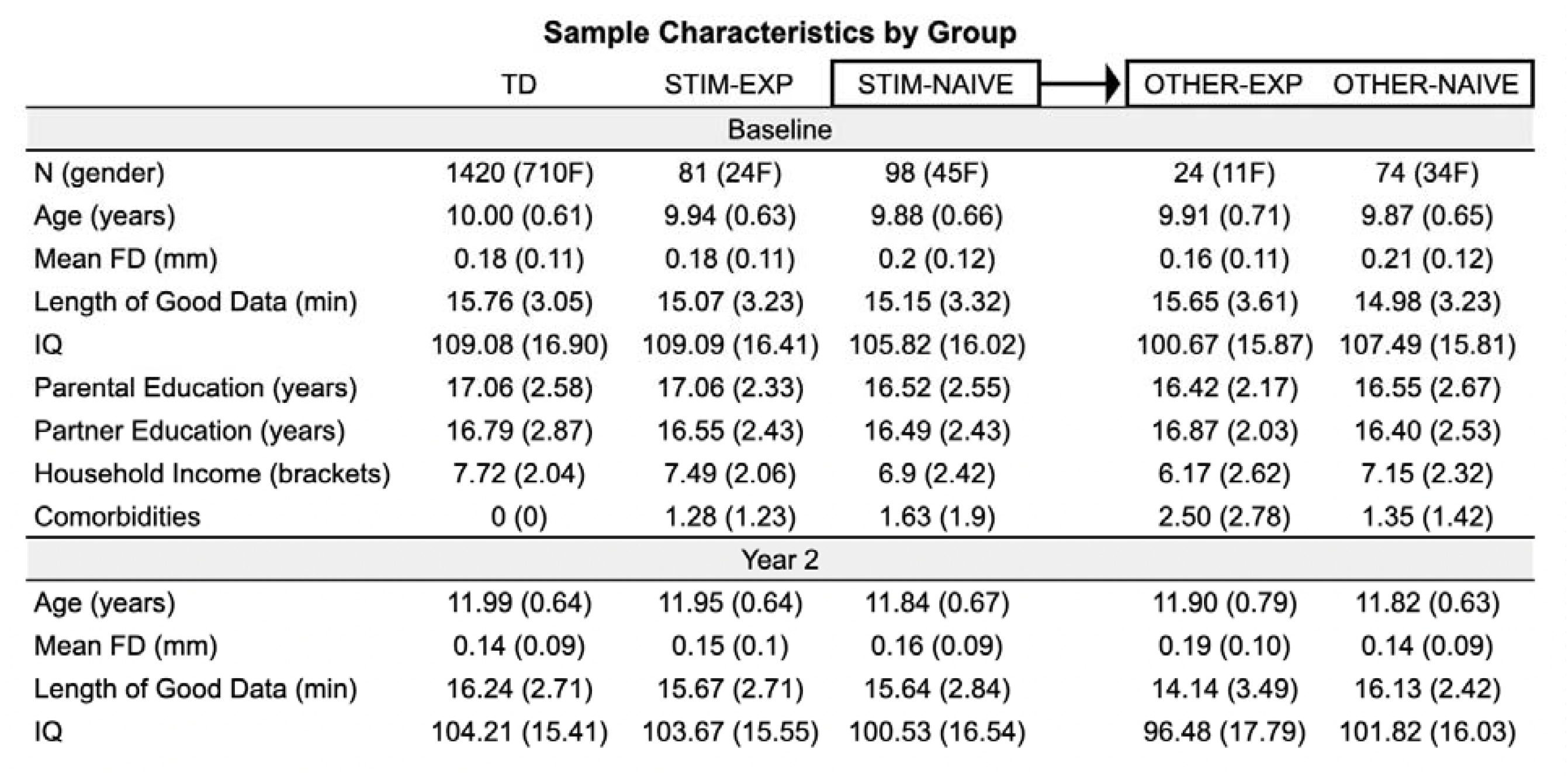
Sample Characteristics of Study Participants and Group Differences. This table displays the means and standard deviations of TD, STIM-EXP, STIM-NAIVE, OTHER-EXP, and OTHER-NAIVE across demographic variables.

An individual was characterized as being stimulant-exposed (STIM-EXP), as compared to stimulant-naive (STIM-NAIVE), if they were reported by their parent or guardian to be using a stimulant at one timepoint or more, whether baseline, the one-year follow-up, or Y2 (a breakdown of all psychotropic medications is reported in Supplemental Material [SM] in Table S1). An individual from the STIM-NAIVE group was characterized as being other-psychotropic medication exposed (OTHER-EXP), as compared to other-naive (OTHER-NAIVE), in the same way. Independent samples t-tests revealed that STIM-EXP (n=81) and STIM-NAIVE (n=98) did not differ on age, IQ (age-corrected picture vocabulary scores from the NIH toolbox), socioeconomic status (SES; averaged z-scores for parental and partner education, coded as the total number of years in school, and combined family income, coded categorically from 1 to 10), number of comorbid diagnoses, or mean FD as well as the length of resting-state data satisfying motion criteria at baseline and Y2 (ps > 0.09) (see Imaging Procedure below). Boys were significantly overrepresented in STIM-EXP ( (1)=4.3, p<0.05). OTHER-EXP (n=24) and OTHER-NAIVE (n=74) did not differ on age, IQ, SES, number of comorbid diagnoses, gender, or mean FD at baseline and Y2 (ps > 0.05).

Our primary outcome measure for symptom improvement was defined as demonstrating reliable T-score reduction on the DSM-oriented ADHD Problems Scale of the CBCL between baseline and Y2. Whether or not decreased T-scores reflected reliable symptom reduction was determined using the reliable change index (RCI), which assesses the reliability of change by taking into account the standard error of measurement: (X at time 2 – X at time 1)/standard error of measurement (36). Applying this method, a difference score (Y2 – baseline) of -11 or lower was considered reliable based on the 80% RCI confidence bound (as in 37), and was used to divide the sample into improvers and no-improvers. Reliable improvers (n=39) did not differ from non-improvers (n=140) on age, IQ, SES, number of comorbid diagnoses, gender, exposure to non-stimulant psychotropic medications, or mean FD as well as length of resting-state data satisfying motion criteria at baseline and Y2 (ps > 0.07). Improvers had significantly higher T-scores on the ADHD Problems Scale at baseline (t(60.6)=3.8, p<0.001) and lower T-scores at Y2 (t(111.6)=-11.1, p<0.001).

Finally, we defined a typically developing (TD) group for comparison with STIM-EXP and STIM-NAIVE. Starting with the same sample of n=11878, we limited inclusion to children who met the following criteria: 1) no diagnoses at baseline based on KSADS-5 (n=9135 out of 11878), 2) T-score < 60 on all DSM-oriented Scales of the CBCL at baseline (n=2303 out of 9135), 3) ABCD Study recommended rs-FC inclusion criteria based on head motion and the quality of FreeSurfer reconstruction at baseline and Y2 timepoints (n=1536 out of 2303), and 4) mean framewise displacement (FD) less than 0.5mm for both baseline and Y2 resting-state data (n=1420 out of 1536). These criteria resulted in a sample of 1420 individuals (mean age at baseline=10.00 years, at Y2=11.99 years; 710 Female), which is described in Table 1.

### Imaging Procedure & Pre-analysis

Children completed four 5-minute runs of resting-state scans on either Siemens or General Electric 3T scanners while they kept their eyes open and focused on a fixation cross (see 14,35,38 for ABCD protocol). While stimulant last use data was collected, this information for baseline and Y2 scanning sessions was missing for those in the present sample, and therefore could not be taken into account (see Discussion). Preprocessing included the removal of initial frames, normalization of voxel time-series, nuisance regression (6 motion parameters and their squares, signal from cerebral white matter, ventricles, and the whole brain, as well as their derivatives), temporal filtering for respiratory signal, and band-pass filtering (0.009-0.08 Hz) (see 35 for details). The average time-series for parcels in the Gordon atlas (39) and subcortical regions from the Freesurfer Aseg Atlas (40) were then correlated to calculate rs-FC, followed by Fisher-z transformation. Each parcel in the Gordon atlas is affiliated with one of 12 functional cortical networks (auditory, cingulo-opercular, cinguloparietal, default mode, dorsal attention, frontoparietal, retrosplenial, salience, somatomotor hand, somatomotor mouth, ventral attention, and visual). Rs-FC values between subcortical regions and parcels were averaged by cortical network, producing a rs-FC value for each subcortical region-network pair. Using these pretabulated data, we selected six striatal regions (left and right caudate, putamen, and nucleus accumbens) as seeds in order to focus on striatal seed-functional network pairs. Given mixed terminology and unclear evidence regarding the functional differentiation of the salience and cingulo-opercular networks (41), we averaged them. Similarly, we averaged the somatomotor hand and mouth networks. Volumes with FD > 0.2mm were excluded from rs-FC calculations, as were volumes with fewer than five contiguous timepoints where FD < 0.2mm for each volume, in order to minimize the effects of motion. Rs-FC was averaged across resting-state runs, weighted by the number of volumes in each run after motion censoring and excluding runs with fewer than 100 volumes (in the retained sample, the baseline range of total volumes was 546-1750; the Y2 range of total volumes was 514-1492). Lastly, we calculated the difference between striatal seed-to-network rs-FC at Y2 and baseline.

### Statistical Analysis

Analyses were conducted in three steps (see Figure 1). For all steps, we ran Bayesian hierarchical regressions (30) with random effects for ABCD site. Bayesian analyses were conducted with the brms package (42) in R and RStudio (version 4.2.2). We used weakly informative priors since we had limited information about the strength of possible associations but did not want to opt for noninformative flat priors in an effort to reduce type I error rates (43). Markov Chain Monte Carlo methods were used to estimate posterior distributions for model parameters with 4 independent chains of 10,000 iterations (2,000 for warm-up). The mean, standard deviation, and two-sided 95% credible interval (CI) of each estimate were used to interpret the findings. Seed-to-network connections selected for interpretation were those whose 95% CIs did not overlap with zero (i.e., differed significantly from zero at the 5% level; as in (44)).

**Figure 1.**
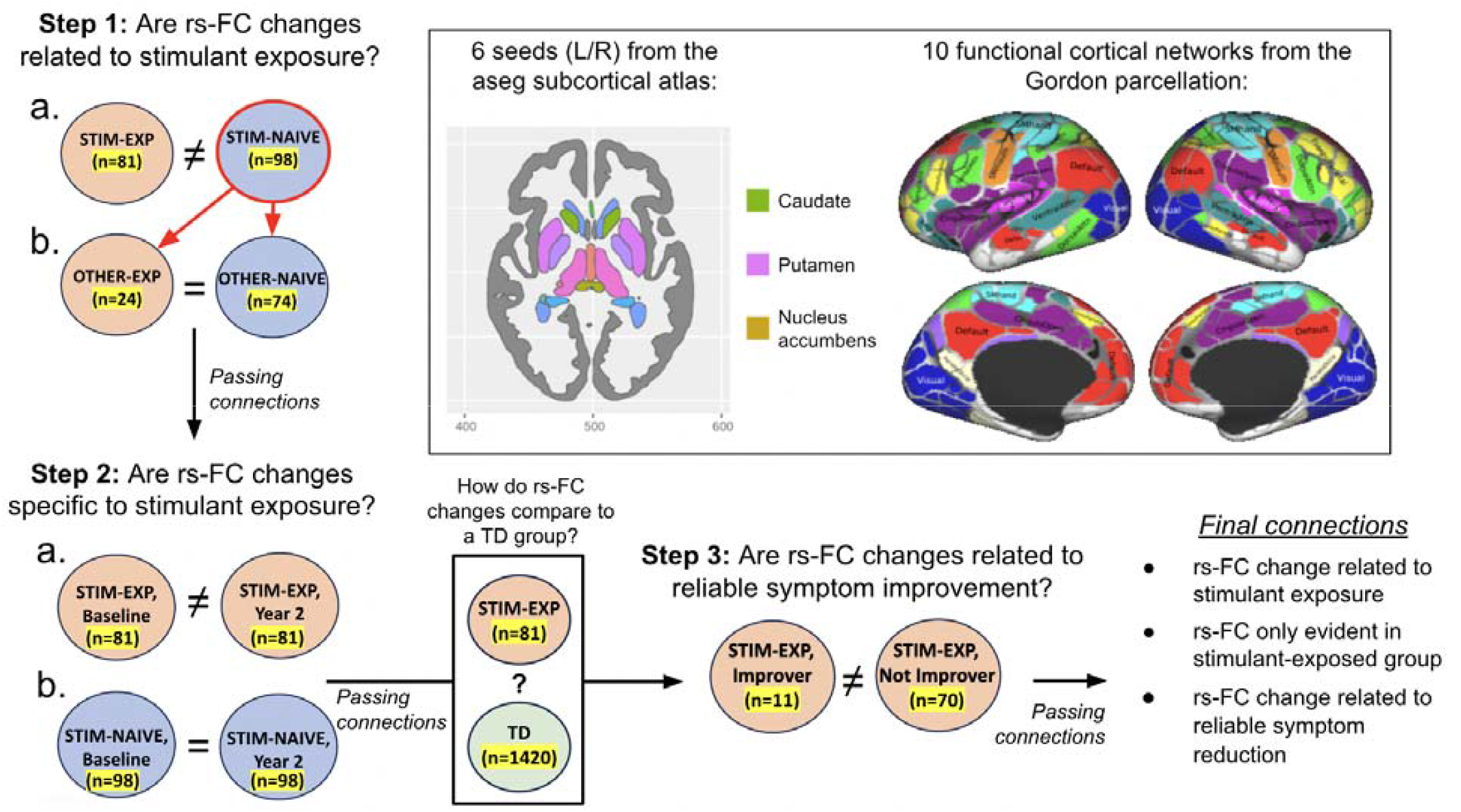
Flowchart for the Three Analytic Steps. We measured rs-FC change between 6 striatal seeds and 10 cortical functional networks from the Gordon parcellation (upper right). First, we assessed evidence for a relationship between rs-FC change and stimulant exposure; second, we confirmed the specificity of rs-FC change to STIM-EXP; and third, we tested associations between rs-FC change and reliable ADHD symptom improvement. Automatic segmentation (Aseg) subcortical atlas from Freesurfer and Gordon parcellation images adapted from https://ggseg.github.io/ggsegExtra/articles/createaseg.html and https://balsa.wustl.edu/WK71 respectively. Rs-FC, resting-state functional connectivity; STIM-EXP, stimulant exposed; STIM-NAIVE, stimulant naive; OTHER-EXP, other psychotropic medication exposed but naive to stimulants; OTHER-NAIVE, completely naive of psychotropic medications; TD, typically developing.

#### Step 1: Predicting Stimulant and Other Psychotropic Medication Exposure

To examine whether change in striatal seed-network rs-FC between baseline and Y2 predicted stimulant exposure (1 for STIM-EXP, 0 for STIM-NAIVE), we ran six Bayesian hierarchical logistic regression models, one for each striatal seed. Each model had 10 total striatal seed-network rs-FC predictors, one for each functional network, and 12 covariates as follows: 4 measures of data quality (mean FD and the square root of the length of rs-FC data after volumes exceeding 0.2mm FD were removed, at baseline as well as Y2), ADHD symptoms at baseline and Y2, a combined measure of SES as it may determine treatment access or intention, a binary measure of exposure to other psychotropic medications, the number of psychiatric comorbidities, IQ at baseline and Y2, and gender, since there was a disproportionate number of boys in STIM-EXP (see results). Next, to ensure that observed associations were specific to simulants, we fit the same logistic regression models to predict other psychotropic medication exposure, by including only participants classified as STIM-NAIVE (1 for OTHER-EXP, 0 for OTHER-NAIVE). Connections identified by both sets of models were dropped from further analysis as they were not selective to stimulant exposure.

#### Step 2: Verifying Driver of Association Between Stimulant Exposure and rs-FC Change; Comparison to Stimulant Naive and TD Groups

For the connections identified in Step 1, we tested for interactions indicating a significant difference in rs-FC across time for STIM-EXP but not for STIM-NAIVE with a Bayesian hierarchical regression model. We arranged data in long format by time as well as by connection and predicted rs-FC with a 3-way interaction between stimulant exposure, time (1 for Y2, 0 for baseline), and connection (i.e., connections passing step 1), controlling for identical covariates to those in step 1. For connections with 3-way interactions significant at the 5% level, we ran post hoc paired samples t-tests, testing rs-FC differences across time for each group, to verify that STIM-EXP was driving the effect (i.e., that the differences in rs-FC across time for connections implicated in the Bayesian model were significant for STIM-EXP and not for STIM-NAIVE). Such connections passed to Step 3.

Lastly, also for all connections identified in Step 1, we tested for 3-way interactions indicating a significant difference in rs-FC across time for STIM-EXP but not for the TD group. For connections with 3-way interactions significant at the 5% level, we again ran post hoc paired samples t-tests, testing rs-FC differences across time for each group, to verify that STIM-EXP was driving the effect.

#### Step 3: Testing Associations Between rs-FC Change and Symptom Change

Finally, we conducted an additional Bayesian hierarchical logistic regression in STIM-EXP to test associations between reliable ADHD symptom improvement based on RCI (defined in Participants section) and rs-FC change of connections passing step 2. Covariates in this model were identical to those in steps 1 and 2, with the addition of length of stimulant exposure (1, 2, or 3 timepoints). As a control analysis, which is presented in the SM, we ran the above model predicting reliable symptom improvement for STIM-NAIVE.

## RESULTS

### ADHD Symptom Change Over Time

Using the conservative RCI criterion, an individual was classified as showing reliable symptom change if the difference between their CBCL ADHD Problems scores at Y2 and baseline was T <= -11. By this criterion, 11 (6 male, 5 female) STIM-EXP improved reliably and 70 (51 male, 19 female) did not, while 28 (13 male, 15 female) STIM-NAIVE improved reliably and 70 did not (40 male, 30 female). The chi-squared test revealed a significant negative association between stimulant exposure and reliable improvement ( (1)=5.0, p<0.05), indicating that stimulant exposure was less likely to be accompanied by reliable symptom improvement. An analysis of continuous symptom scores by stimulant group and time conducted for descriptive purposes (see SM “Change in ADHD Symptom Scores” for details) clarified that while mean ADHD symptoms improved for both groups, STIM-NAIVE improved to a greater extent than STIM-EXP (see SM Figure S1-note the high individual variation in change from baseline to Y2). Additionally, since applying exclusion criteria for head motion inevitably results in an unrepresentative sample (i.e., children who can stay still), we removed these criteria as well as the criterion of ADHD Problems T-score > 60 at baseline in order to look at continuous symptom scores by stimulant group and time with the most lenient criteria (i.e., only ADHD on KSADS at baseline). In this larger sample (n=556), by the above RCI criterion, 32 (23 male, 9 female) STIM-EXP improved reliably and 200 (152 male, 45 female, 3 missing) did not, while 54 (31 male, 22 female, 1 missing) STIM-NAIVE improved reliably and 270 (182 male, 83 female, 5 missing) did not. The chi-squared test revealed no association between stimulant exposure and reliable improvement (χ^2^ (1)=0.65, p=0.42). In a continuous analysis of symptoms for this larger sample, mean ADHD symptoms again improved for both STIM-EXP and STIM-NAIVE across time, and again STIM-NAIVE improved to a greater extent (see SM Figure S2). An analysis of reliable symptom improvement for OTHER-EXP and OTHER-NAIVE within the STIM-NAIVE group is also presented in SM (see “ADHD Symptom Change for OTHER-EXP and OTHER-NAIVE”), which did not reveal an association between other medication exposure and reliable symptom improvement.

### Stimulant Exposure and Change in Striatal rs-FC: Steps 1 and 2

Results from step 1 revealed strong evidence for associations between stimulant exposure and change in nine striatal-cortical functional connections (Figure 2a): left caudate and cingulo-opercular/salience network (Est.=5.02, sd=1.84, 95% CI=[1.53,8.74], ESS=25410, Rhat=1.00), left caudate and frontoparietal network (Est.=-8.84, sd=2.68, 95% CI=[-14.24,-3.75], ESS=21531, Rhat=1.00), left putamen and cinguloparietal network (Est.=2.77, sd=1.14, 95% CI=[0.59,5.08], ESS=31804, Rhat=1.00), left putamen and frontoparietal network (Est.=-3.46, sd=1.26, 95% CI=[-6.02,-1.08], ESS=19853, Rhat=1.00), left putamen and somatomotor network (Est.=6.38, sd=2.58, 95% CI=[1.57,11.69], ESS=27842, Rhat=1.00), left nucleus accumbens and dorsal attention network (Est.=-4.99, sd=2.61, 95% CI=[-10.22,-0.03], ESS=26934, Rhat=1.00), right putamen and dorsal attention network (Est.=-3.31, sd=1.59, 95% CI=[-6.49,-0.24], ESS=18542, Rhat=1.00), right putamen and ventral attention network (Est.=3.53, sd=1.55, 95% CI=[0.56,6.63], ESS=25393, Rhat=1.00), and right putamen and visual network (Est.=-3.46, sd=1.31, 95% CI=[-6.12,-0.97], ESS=20782, Rhat=1.00).

**Figure 2.**
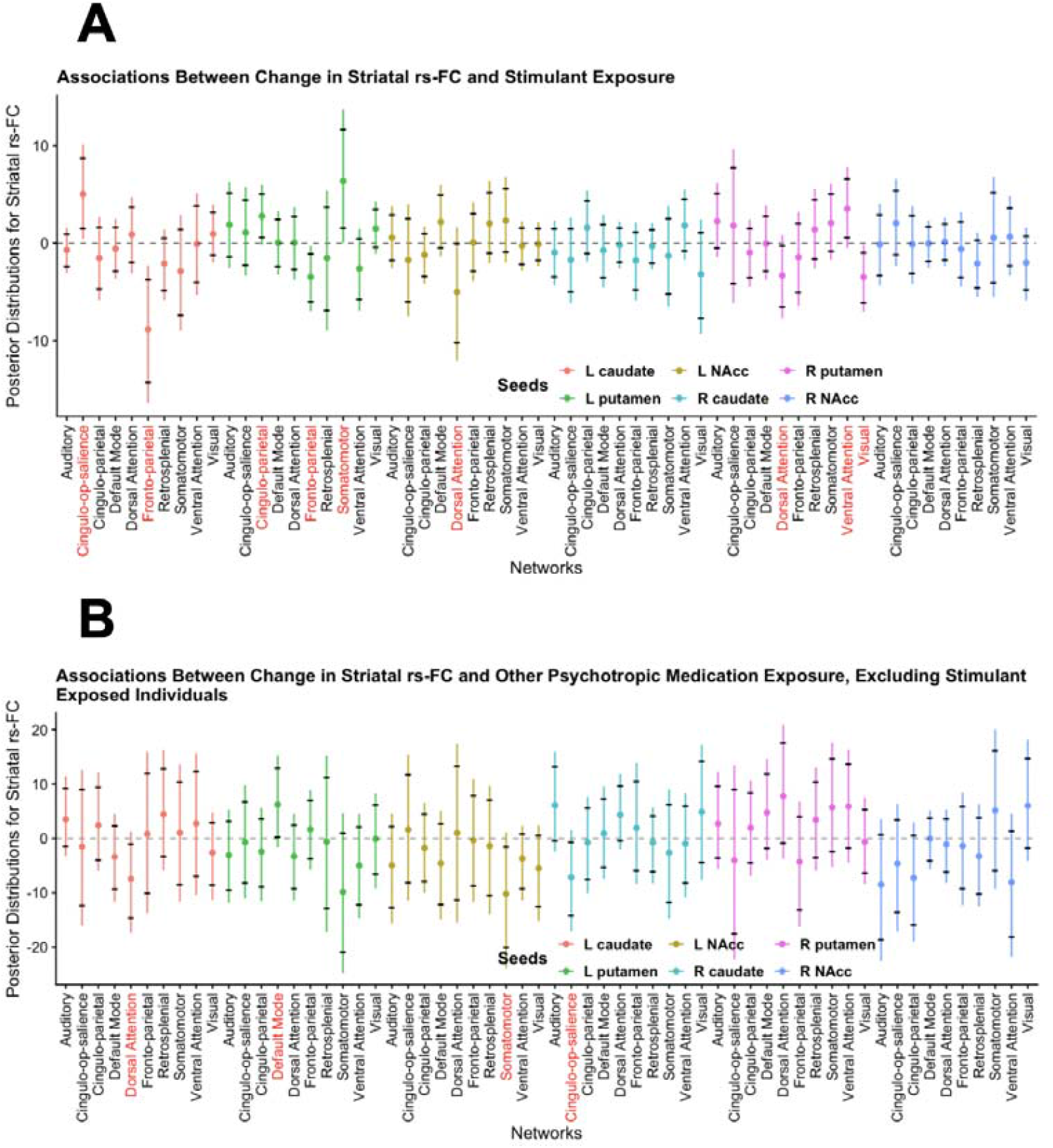
Results of Bayesian Hierarchical Logistic Regression Models Estimating the Association Between rs-FC Change and **(a)** Stimulant Exposure and **(b)** Other Psychotropic Medication Exposure. This figure shows estimates on the y-axes of the relationships between change in rs-FC from baseline to year 2 and medication exposure, which is stimulant exposure in part a and other psychotropic medication exposure in part b, which excludes participants exposed to stimulants. Striatal seeds are shown in different colors and cortical functional networks are displayed along the x-axes. The bars span 99% of the estimates for each association and the tick marks reflect the 95% credible intervals. Associations where the 95% credible interval does not overlap with 0 are highlighted on the x-axes with red font for the implicated cortical functional network. Rs-FC, resting-state functional connectivity.

A nonoverlapping set of connections was modulated by exposure to non-stimulant medications. Results revealed strong evidence for associations between other psychotropic medication exposure and change in four striatal-cortical functional connections (Figure 2b): left caudate and dorsal attention network (Est.=-7.48, sd=3.48, 95% CI=[-14.63,-1.00], ESS=26521, Rhat=1.00), left putamen and default mode network (Est.=6.20, sd=3.20, 95% CI=[0.25,12.92], ESS= 24954, Rhat=1.00), left nucleus accumbens and somatomotor network (Est.=-10.20, sd=4.73, 95% CI=[-20.08, 1.51], ESS=18934, Rhat=1.00), and right caudate and cingulo-opercular/salience network (Est.=-7.18, sd=3.48, 95% CI=[-14.22,-0.66], ESS=29944, Rhat=1.00). Connections identified by the two sets of models did not overlap, suggesting that the nine connections sensitive to stimulant exposure were not also sensitive to psychotropic medications generally.

In step 2, we further examined the selectivity of sensitivity to stimulant exposure of the nine connections identified in step 1. Results revealed three connections where there was evidence for the relationship between time and rs-FC differing between STIM-EXP and STIM-NAIVE: left caudate and frontoparietal network (Est.=-0.07, sd=0.04, 95% CI=[-0.14,-0.00], ESS=6649, Rhat=1.00), left putamen and frontoparietal network (Est.=-0.10, sd=0.04, 95% CI=[-0.17,-0.03], ESS=6744, Rhat=1.00), and right putamen and visual network (Est.=-0.09, sd=0.04, 95% CI=[-0.16,-0.02], ESS=6443, Rhat=1.00). Post hoc paired samples t-tests (see Table 2) comparing rs-FC in baseline and Y2, for STIM-EXP and STIM-NAIVE separately, revealed that for each connection, rs-FC significantly decreased across time for STIM-EXP but did not change for STIM-NAIVE (Figure 3). These three connections therefore passed to step 3, to test for association with reliable symptom improvement.

**Figure 3.**
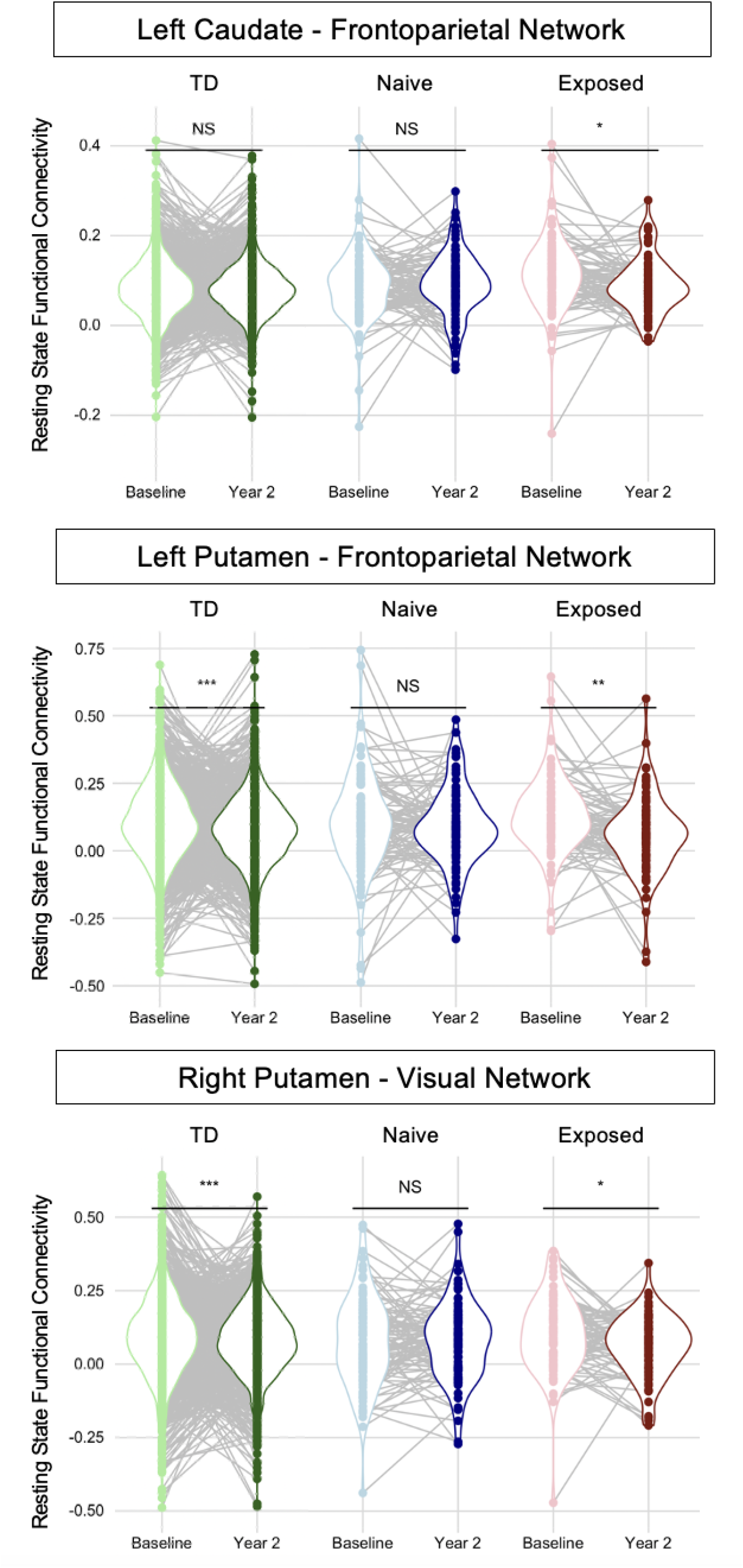
Rs-FC at Baseline and Year 2 Timepoints for Connections Passing Analysis Steps 1 and 2, Separately for Stimulant Exposure Groups and the TD Group. This figure shows rs-FC change for the connections where 1) there was strong evidence for an association between rs-FC change and stimulant exposure, and 2) there was strong evidence for a stimulant exposure X time X connection interaction. We verified that rs-FC change was driven by STIM-EXP with paired samples t-tests (asterisks reflecting p-values are displayed above the violin plots). Rs-FC decreased in STIM-EXP for all identified connections, such that positive rs-FC became less positive for STIM-EXP and not for STIM-NAIVE. Decreased positive rs-FC was also evident in the TD group for two connections, left putamen – frontoparietal network and right putamen – visual network. Individual participants’ rs-FC is reflected as dots which are connected across time. TD, typically developing; * p < 0.05; ** p < 0.005; *** p < 0.001.

**Table 2.**
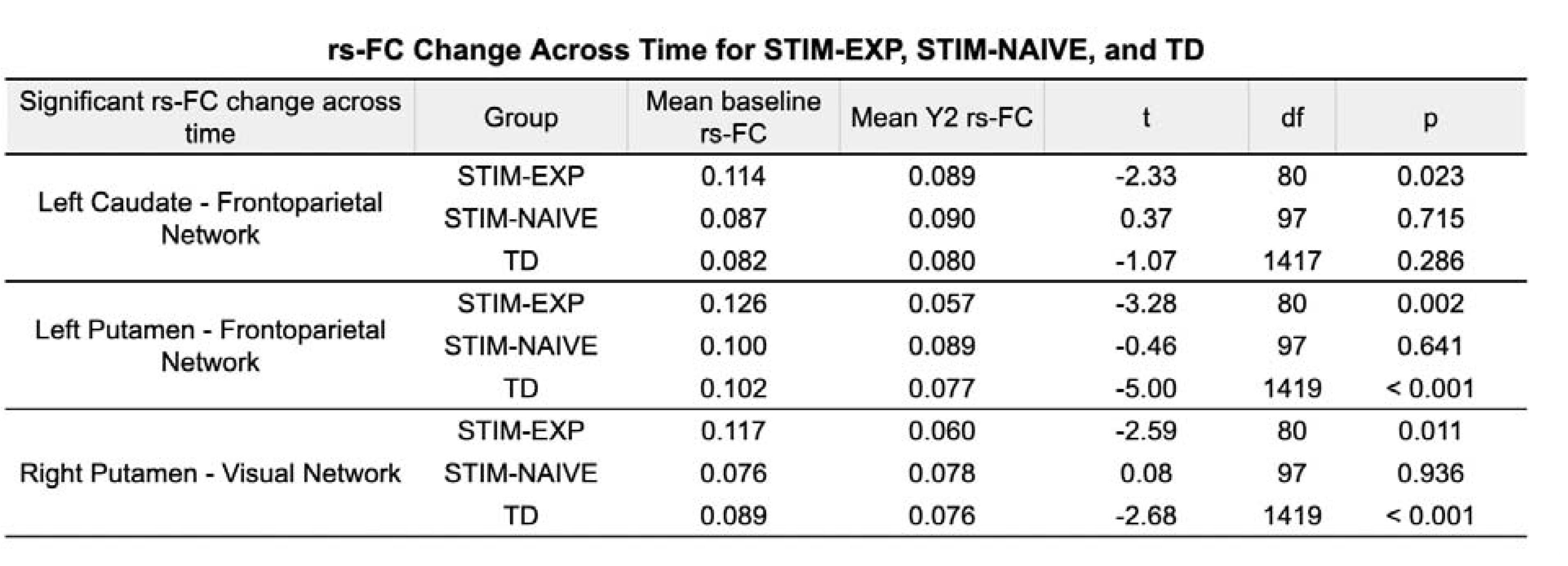
Rs-FC Change Across Time. This table displays the mean rs-FC at baseline and year 2 for each connection with any evidence of rs-FC change differing by group in Step 2. Post hoc paired samples t-tests are displayed for each group testing whether rs-FC differed between baseline and year 2. Rs-FC, resting-state functional connectivity; STIM-EXP, stimulant exposed; STIM-NAIVE, stimulant naive; TD, typically developing; Y2, year 2.

Lastly, to determine whether those changing connections for STIM-EXP showed a normalizing trajectory, we ran an additional Bayesian hierarchical regression model to further test the selectivity of sensitivity to stimulant exposure of the nine connections identified in step 1 by comparing STIM-EXP and the TD group. Results revealed that for two connections there was evidence for the relationship between time and rs-FC differing between STIM-EXP and TD. Both connections were already identified in the previous analysis: left putamen and frontoparietal network (Est.=0.05, sd=0.03, 95% CI=[0.00,0.10], ESS=6852, Rhat=1.00) and right putamen and visual network (Est.=0.05, sd=0.03, 95% CI=[0.01,0.10], ESS=6693, Rhat=1.01). Post hoc paired samples t-tests (see Table 2) comparing rs-FC in baseline and Y2, for STIM-EXP and the TD group separately, revealed that for both connections, rs-FC significantly decreased across time for both STIM-EXP and the TD group, and that the magnitude of reduction was greater for STIM-EXP than for TD, suggesting a potential normalizing effect. An identical Bayesian hierarchical regression model comparing STIM-NAIVE and the TD group found no evidence for rs-FC change across time differing by group for any of the nine connections.

### Associations with Symptom Improvement: Step 3

Of the three connections where change in rs-FC was driven by stimulant exposure, only right putamen-visual network showed strong evidence for an association between rs-FC change and the likelihood of reliable symptom improvement based on RCI in STIM-EXP (Est.=-12.54, sd=5.79, 95% CI=[-25.07,-2.55], ESS=8210, Rhat=1.00) (Figure 4), indicating that greater reduction in rs-FC was linked to higher likelihood of reliable symptom improvement. When we repeated the above model for STIM-NAIVE, there was no evidence for any associations between rs-FC and reliable symptom improvement (reported in SM). This result was expected, since connections were selected for not demonstrating change at the group level for STIM-NAIVE, but we wanted to confirm the specificity of the results to STIM-EXP.

**Figure 4.**
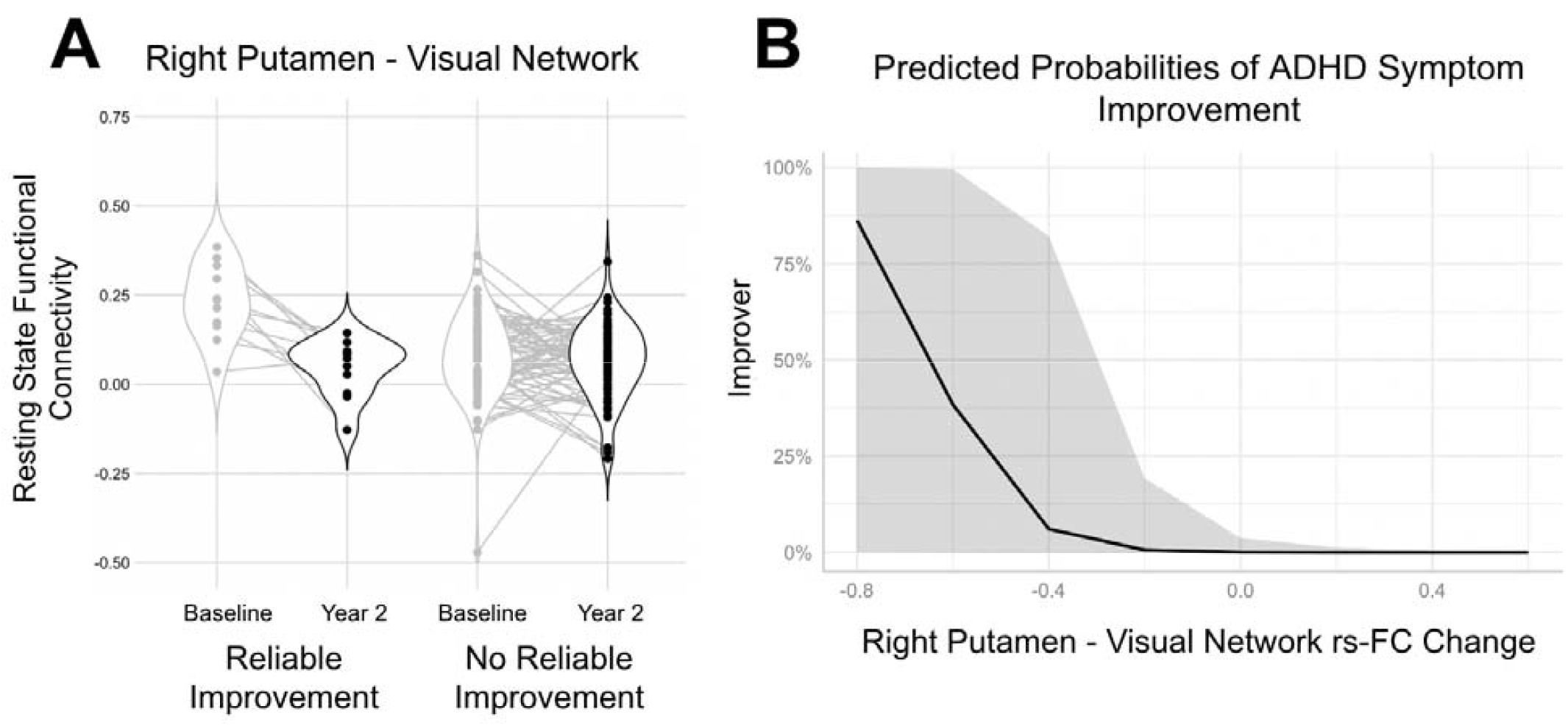
Right Putamen-Visual Network rs-FC Change for Reliable Improvement and No Reliable Improvement Groups. A) violin plots for rs-FC at baseline and year 2, excluding stimulant-naive participants. Individual participants’ rs-FC values are reflected as dots connected across time. B) Predicted probabilities plot for rs-FC change (Y2 – baseline) showing probability of having reliable improvement, along with the 95% confidence interval.

## DISCUSSION

We examined change in rs-FC between striatal regions and canonical cortical functional networks which was associated with exposure to stimulant medication over a 2-year period in preadolescent children with ADHD in the ABCD dataset. Results revealed nine striatal-cortical functional networks where change in rs-FC was associated with exposure to stimulant medication, but not to other psychotropic medications. The identified connections were primarily between left hemisphere seeds and canonical executive networks, including left caudate and cingulo-opercular/salience as well as frontoparietal networks; left putamen and cinguloparietal, frontoparietal, and somatomotor networks; and left nucleus accumbens and dorsal attention network. Three connections were identified with a right hemisphere seed, right putamen, two with canonical executive networks, dorsal as well as ventral attention networks, and another with a sensory network, the visual network. Of these nine connections, left caudate-frontoparietal network, left putamen-frontoparietal network, and right putamen-visual network demonstrated rs-FC change over the 2 years in children exposed to stimulant medication and not in those who were stimulant naive. Further, left putamen-frontoparietal network and right putamen-visual network demonstrated stronger rs-FC reduction in children exposed to stimulant medication when compared to a group of typically developing children without ADHD, who also had reduced rs-FC across time for these two connections, suggesting a normalizing effect of stimulant exposure. Of the three connections differing between stimulant exposed and naive children with ADHD, only rs-FC change of right putamen-visual network was associated with reliable symptom improvement, defined as having a T-score reduction on the ADHD Problems Scale of the CBCL (Y2 – baseline) of -11 or lower, based on the 80% reliable change index confidence bound. Together, these findings indicate widespread modulation of striatal functional connectivity over 2 years at the cusp of adolescence that distinguished stimulant exposure in children with ADHD, but only rs-FC of the putamen and visual network was related to reliable symptom improvement, such that greater rs-FC reduction was associated with a greater likelihood of improving across time.

There are several factors to keep in mind for interpreting the current findings. First, the amount of medication exposure cannot be precisely measured as the ABCD protocol includes only the parent’s report at the time of the baseline and Y2 MRI scans and the year 1 interim visit, and no monitoring of medication status during the year. Classification of a participant as stimulant exposed was based on taking a stimulant at *any* of the three visits with the rationale that it approximated real-world treatment conditions, which are marked by variation in dose, lack of consistent medication compliance, and intermittent exposure. Exposure to other psychotropic medications followed the same rationale. Second, several factors that are relevant to functional brain variation were controlled by including as covariates, namely head motion/data quality (two measures: mean FD and the square root of the length of rs-FC data following removal of frames exceeding 0.2mm FD), ADHD symptoms, socioeconomic status (composite defined by parental and partner education as well as combined family income), gender, IQ, presence of other psychotropic medications, and number of psychiatric comorbidities. However, as patterns of comorbidity are not stable across time and fluctuations in symptoms are not linear (45), the current results should not be extrapolated to beyond this 2-year window in early adolescence. Third, stimulant washout information for baseline and Y2 scanning sessions was missing for all subjects in the present sample and could not be controlled for, making any acute effects of stimulant treatment or withdrawal uncharacterizable. Nonetheless, recent work with the ABCD sample has found no effect of a 24-48 stimulant washout period on functional network rs-FC (examining default mode and frontoparietal networks) in ADHD, with comparison to stimulant exposed children, assuaging concerns that this could be confounding (46). Fourth, an inherent limitation of large-scale rs-FC networks is that they represent averages across many parcels, resulting in lost information at the parcel-level. However, our sample of children with ADHD at the baseline timepoint was underpowered to look at relationships among several hundred parcels, making a focus on the whole network an efficient dimensionality reduction strategy. It is also an open question what functional unit or units are best for explaining variation in symptoms, or which would be sensitive to stimulant exposure. Moreover, there is high functional variation across brains such that a parcel-level analysis may be best suited for a precision or individualized approach. Fifth, as an additional dimensionality reduction step we averaged some functional networks (i.e., cingulo-opercular and salience networks; and somatomotor hand and somatomotor mouth networks) based on theoretical views that there may not be a meaningful distinction for the purposes of the present analysis (41). Sixth, stimulant and other psychotropic medication exposure was based on parent report, which could be incorrect. Seventh, since we required high quality imaging data, the sample is not representative of all children with ADHD, but to a subset who were able to remain still.

ADHD symptom change was evident in both STIM-EXP and STIM-NAIVE groups, and therefore was not selective to stimulant exposure. Binarized symptom improvement was defined as demonstrating reliable T-score reduction (-11 or less, based on the reliable change index) on the DSM-oriented ADHD Problems Scale of the CBCL. Our criterion was a conservative definition of symptom improvement, since we wanted to capture change that was reliable and clinically meaningful. While this strengthens the rigor of our improvement classification, it is also important to note that considerable stability across time has been found for the CBCL in kids with ADHD, validating it as a longitudinal measure (47,48). A greater proportion of children from STIM-NAIVE compared to STIM-EXP showed reliable symptom improvement when considered under the conservative inclusion criteria (KSADS + CBCL + imaging quality). While this may appear surprising, it underscores the noted heterogeneity in treatment efficacy in ADHD, with 20-35% of people having inadequate responses to treatment with stimulants in clinical trials (49). On analysis of mean differences, while both groups had lower scores at Y2 than at baseline, STIM-NAIVE showed greater reduction. This result cannot be explained by an unrepresentative sample (e.g., due to restrictive head motion inclusion criteria), as the result was replicated in a sample selected with lenient inclusion criteria (ADHD at baseline based on KSADS only; n=556). Reliable symptom improvement for STIM-NAIVE was also not driven by exposure to other psychotropic medications, since when STIM-NAIVE was split into OTHER-EXP and OTHER-NAIVE, there were no differences in reliable symptom improvement. These contrasting results between STIM-EXP and STIM-NAIVE could be theoretically explained by a bias in who is prescribed stimulant medications (i.e., a patient selection bias), as previous work has shown that ADHD symptom severity is a predictor of stimulant use (50), and more severe symptoms may be more resistant to change. While such a bias may exist, STIM-EXP and STIM-NAIVE did not differ on mean ADHD symptoms at baseline in the present sample. Boys were more likely to be prescribed stimulants, and are also known to present with more externalizing symptoms (51), which could have biased symptom results if the CBCL ADHD Problems Scale was primarily picking up externalizing problems. However, the inclusion of gender as a covariate means it did not drive the main results. Alternatively, the observation of less symptom improvement over time in STIM-EXP may reflect maladaptive behaviors that were exacerbated by medication (e.g., irritability, mood disturbances), which are particularly noted in amphetamine-derived stimulants (52). Therefore, while core symptoms of ADHD were reduced by stimulant medication, other behaviors may have worsened (e.g., emotional lability, see 53), which the ADHD Problems Scale of the CBCL was likely not sufficiently sensitive to parse. Comprehensive characterization of ADHD symptoms was not part of the ABCD study design and therefore our present conclusions are limited to the CBCL ADHD Problems Scale. In accordance with the present findings, extended use of stimulant medication in childhood is not always found to be linked to long-term reduction in symptom severity (54), and is sometimes only marginally associated with improvements in adult outcomes, such as protection against later substance abuse (55) and school absenteeism (56), without a strong link to long-term ADHD symptom outcomes. This may in part be due to individual heterogeneity in responses to stimulant treatment (e.g., 57), obscuring consistent group level effects. This could also be due to the fact that many studies of symptom progression are prospective follow-up studies with higher potential for selection bias as compared to controlled trials, which in contrast have more consistently shown symptom improvement across time for stimulant-treated individuals (58). Since the current study capitalized on a naturalistic design, it may not be surprising to see a weaker trajectory of improvement for stimulant-exposed compared to stimulant-naive children.

The integration of cortico-striatal functional circuits is crucial to carrying out cognitive and motor processes which are implicated in ADHD and lead to purposeful action (59). The present imaging results indicated that rs-FC of cortico-striatal circuitry was sensitive to chronic stimulant exposure. We observed widespread change in rs-FC between striatal seeds and canonical executive cortical networks (cingulo-opercular/salience, frontoparietal, cinguloparietal, and ventral and dorsal attention networks), circuitry that has also been implicated in findings from studies of acute stimulant exposure (21). These findings extend work suggesting that large-scale dysfunction at the cortical network level may be better able to account for the complex patterns of impairment seen in ADHD compared to more fine-grained loci of dysfunction. Canonical executive networks, for instance, have shown abnormalities in adult ADHD which are partially normalized by acute stimulant exposure (60). Stimulant trials in children and adolescents have also demonstrated modulation of rs-FC at the network-level, often focusing on default mode and canonical executive networks and striatal regions (e.g., 5,19). Complementing such controlled paradigms, we show that rs-FC between large-scale executive networks and striatal regions is sensitive to stimulant exposure in a large naturalistic study, mirroring conditions of the real world. A focus on the network level may also be especially important in the context of developing therapies, such as transcranial magnetic stimulation (TMS). Application of TMS, for instance, to dorsolateral prefrontal cortex for ADHD (61), modulates the entire network, i.e., all other regions with which it is functionally and structurally connected (62). Therefore, identifying how stimulant treatment modulates executive network rs-FC, and its relation to individual symptom trajectories, may lead to new hypotheses about therapeutic targets.

The present findings also extend work characterizing the relationship between stimulant modulation of striatal-cortical network rs-FC and changes in ADHD symptom severity. While previous studies have largely focused on the default mode and frontoparietal networks, our findings suggest that striatal rs-FC with a sensory network, the visual network, may have clinical relevance. Specifically, right putamen-visual network demonstrated reduced rs-FC across time in the stimulant exposed group, while no change was identified in the stimulant naive group. This reduction in rs-FC exceeded in magnitude a similar reduction in a typically developing group, suggesting a normalizing effect of stimulant exposure on rs-FC of right putamen-visual network. Higher putamen-visual network rs-FC has been linked to more severe symptoms of impulsivity and hyperactivity (63), further suggesting that reduced rs-FC indicates a therapeutic and normalizing change. Accordingly, while only 14% (11/81) of stimulant-exposed children showed reliable symptom improvement in the present sample, they were distinguished by greater reduction in right putamen-visual network rs-FC when compared to those without reliable symptom improvement. While an n of 11 is small, by defining symptom improvement taking into account measurement variability of the CBCL DSM-oriented ADHD Scale, we ensured that the symptom improvement was reliable and clinically meaningful, and thereby, rs-FC results interpretable. The visual network broadly has been associated with a wide range of behaviors relevant to ADHD pathology, including visual attention and inattentive symptoms (15,64). Since the putamen is classically linked to motor control, the effects of putamen-visual network rs-FC modulation may relate broadly to attentional problems expressed as motor hyperactivity and restlessness. Finally, left putamen-frontoparietal network also demonstrated reduced rs-FC across time in the stimulant exposed group, with no change identified in the stimulant naive group. This reduction in rs-FC similarly exceeded in magnitude a reduction in a typically developing group, again suggesting a normalizing effect of stimulant exposure. Functional abnormalities have long been noted in ADHD in frontostriatal and frontoparietal circuits, mediated by dopamine function (65,66), however we did not identify a link between this connection and reliable symptom improvement. This could be due in part to symptom change in the present sample being driven by visual and lower-order attentional capacity (67), associated with somatosensory regions, rather than higher-order functions associated with the frontoparietal network. However, we were not able to parse this distinction with the CBCL ADHD Problems Scale.

In sum, our results reveal that stimulant exposure under real-world conditions over two years widely modulates bilateral large-scale cortical network connectivity with the striatum in preadolescent children with ADHD. While the therapeutic significance of stimulant exposure was somewhat limited, our findings highlight the potential clinical relevance of a visuo-motor network. We hope that these findings will generate new hypotheses to test in future randomized controlled trials.

## Data Availability

All data produced in the present study are available upon reasonable request to the authors

https://nda.nih.gov/abcd

## ACKNOWLEDGEMENTS

We would like to acknowledge and thank the ABCD study.

## AUTHOR CONTRIBUTIONS

AK performed the analyses, BH compiled the medication data, and AK, HX and CJV wrote the paper.

