## Supplemental Material for "Change in Striatal Functional Connectivity Networks Across Two Years Due to Stimulant Exposure in Childhood ADHD: Results from the ABCD Sample"

**Breakdown of All Psychotropic Medication Exposure**

Individuals were characterized as being exposed to a psychotropic medication if they were reported by their parent or guardian to be using the medication at the baseline timepoint, the one year follow-up, or Y2 (or a combination of all three). Table S1 describes the reported medication usage for each timepoint for the following classes: ADHD stimulant (Amphetamine, Methylphenidate, Dextroamphetamine, Dexmethylphenidate, Adderall, Adzenys, Concerta, Cotempla, Evekeo, Focalin, Metadate, Quillivant), ADHD non-stimulant (Atomoxetine, Guanfacine, Intuniv, Strattera), antidepressant (Celexa, Citalopram, Desvenlafaxine, Duloxetine, Escitalopram, Fluoxetine, Fluvoxamine, Imipramine, Lexapro, Mirtazapine, Prozac, Remeron, Sertraline, Trazodone, Wellbutrin, Zoloft), anticonvulsant (Divalproex sodium, Gabapentin, Lamotrigine, Neurontin), antihypertensive (Tribenzor, Catapres, Clonidine), and antipsychotic (Abilify, Aripiprazole, Lurasidone, Quetiapine, Risperdal, Risperidone, Seroquel, Ziprasidone). In summary, 60 participants had only ever been exposed to stimulant medication, 21 had been exposed to stimulant medication and at least one other psychotropic medication, 24 had only ever been exposed to a non-stimulant psychotropic medication, and 74 had never been exposed to any medication.

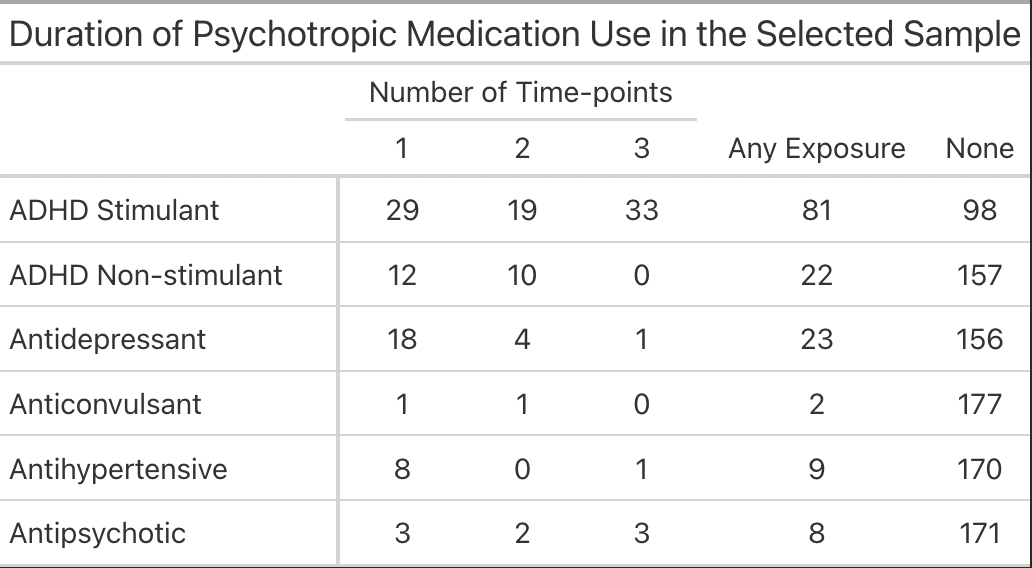

**Table S1**. Breakdown of Psychotropic Medications: Usage and Duration Among Study Participants. This table provides a description of the classes of psychotropic medications used by participants as well as the total number of participants reporting use at any time-point in the “Any Exposure” column. Further, we broke down overall use by the total number of time-points use was reported (across baseline, year 1, and year 2) in order to summarize the duration of use.

**Change in ADHD Symptom Scores**

In addition to examining ADHD symptom scores categorically by whether or not symptom improvement was reliable (i.e., based on the reliable change index), we examined symptom scores continuously, running a 2X3 repeated measures Analysis of Variance (ANOVA) to compare symptom means across time (baseline, Y1, Y2) and stimulant group (STIM-EXP, STIM-NAIVE). Results indicated a significant main effect of time (F(2,349)=31.56, p<0.001), such that ADHD symptoms improved over the 2-year period for both STIM-EXP and STIM-NAIVE groups, as well as a main effect of group (F(1,175)=13.64, p<0.001), such that STIM-EXP had more severe ADHD symptoms overall. There was also a significant time by group interaction effect (F(2,349)=5.54, p=0.004). Post hoc paired samples t-tests, comparing ADHD symptoms at baseline and Y2 for STIM-EXP and STIM-NAIVE separately, indicated that while both groups improved, the magnitude of symptom improvement across the 2-year period was higher for STIM-NAIVE (t(80)=7.6, p<0.001; Cohen’s d=0.88) than it was for STIM-EXP (t(80)=2.8, p<0.01; Cohen’s d=0.37). Additional post hoc t-tests indicated that while there was no group difference in symptom severity between STIM-EXP and STIM-NAIVE at baseline (t(169.04)=1.2, p=0.23; Cohen’s d=0.18), symptoms did significantly differ between STIM-EXP and STIM-NAIVE at Y2 (t(175.19)=4.1, p<0.001; Cohen’s d=0.60), such that STIM-NAIVE was less severely impaired. Change in ADHD symptoms for each group separately is summarized in Figure S1.

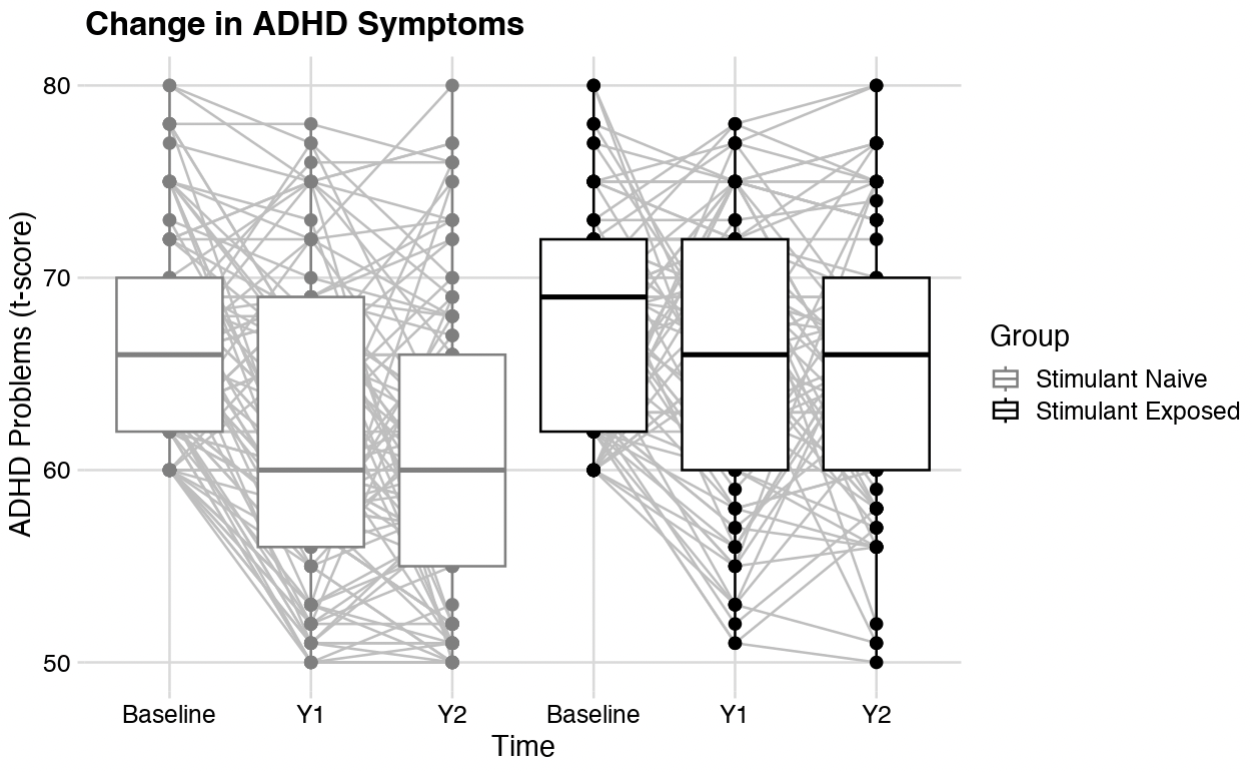

**Figure S1.** T-scores on the ADHD Problems Scale of the CBCL at each Timepoint, Separately for Stimulant Exposed and Naive Groups. Boxplots show the median as well as the 25^th^ and 75^th^ percentiles. Individual participants’ scores are reflected as dots connected across time.

**Change in ADHD Symptom Scores with Lenient Inclusion Criteria**

Since our original inclusion criteria resulted in an inevitably unrepresentative sample (i.e., children with ADHD who could remain still in the MRI), we also examined symptom scores continuously for a larger sample with a lenient inclusion: ADHD diagnosis at baseline only according to KSADS (n=556). We repeated the 2X3 repeated measures Analysis of Variance (ANOVA) to compare symptom means across time (baseline, Y1, Y2) and stimulant group (STIM-EXP, STIM-NAIVE). Results indicated a significant main effect of time (F(2,1095)=51.49, p<0.001), such that ADHD symptoms improved over the 2-year period for both STIM-EXP and STIM-NAIVE groups, as well as a main effect of group (F(1,515)=45.41, p<0.001), such that STIM-EXP had more severe ADHD symptoms overall. There was again a significant time by group interaction effect (F(2,1095)=3.01, p<0.05). Post hoc paired samples t-tests, comparing ADHD symptoms at baseline and Y2 for STIM-EXP and STIM-NAIVE separately, indicated that while both groups improved, the magnitude of symptom improvement across the 2-year period was larger for STIM-NAIVE (t(323)=8.7, p<0.001; Cohen’s d=0.47) than it was for STIM-EXP (t(231)=4.7, p<0.001; Cohen’s d=0.32). Additional post hoc t-tests indicated that STIM-EXP and STIM-NAIVE differed in symptom severity both at baseline (t(491.72)=4.3, p<0.001; Cohen’s d=0.37) and Y2 (t(487.99)=5.9, p<0.001; Cohen’s d=0.51), but that this effect was strongest at Y2. Change in ADHD symptoms for each group separately in the larger sample is summarized in Figure S2.

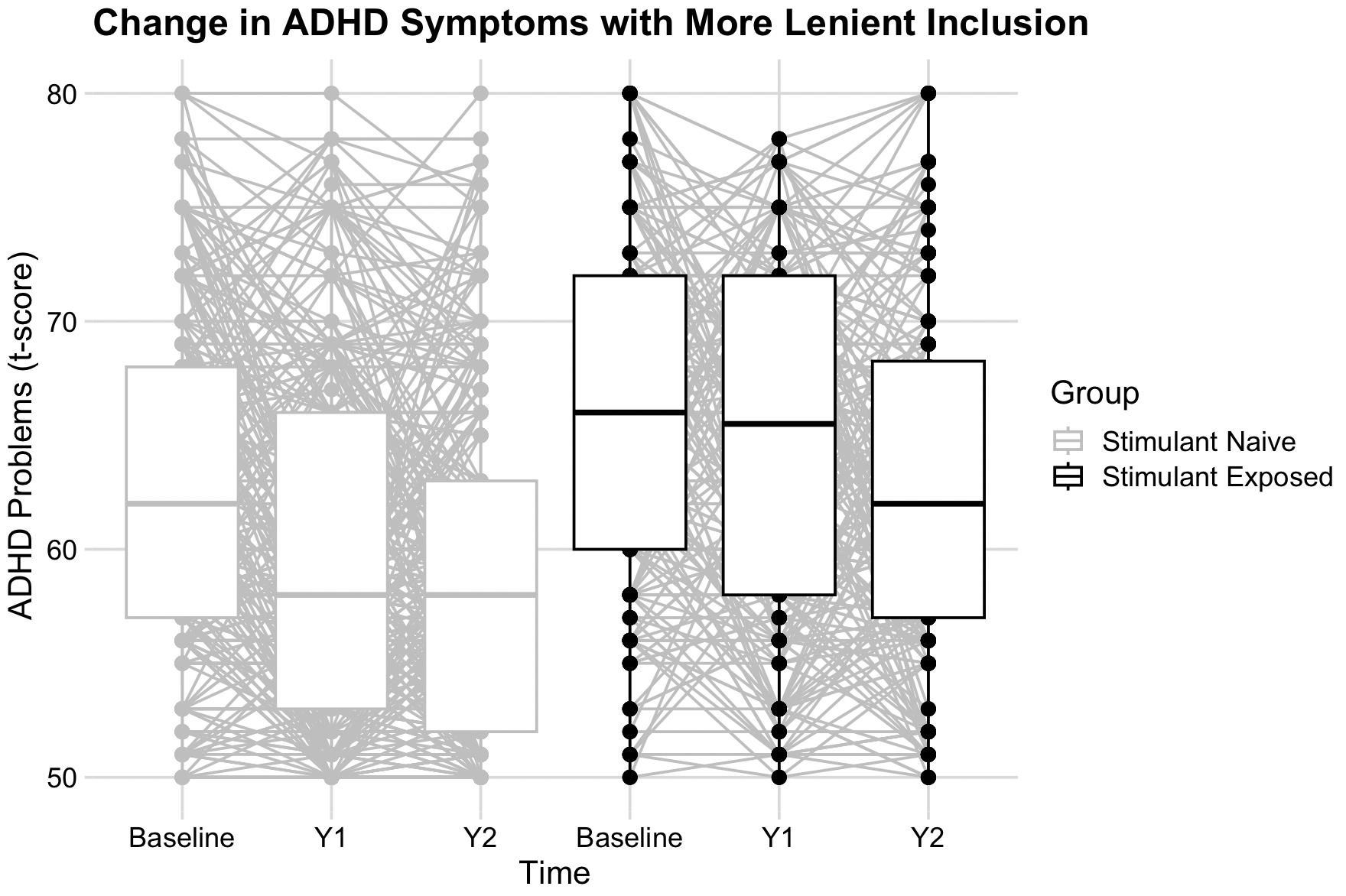

**Figure S2.** T-scores on the ADHD Problems Scale of the CBCL at each Timepoint, Separately for Stimulant Exposed and Naive Groups, in the Larger Sample. Boxplots show the median as well as the 25^th^ and 75^th^ percentiles. Individual participants’ scores are reflected as dots connected across time.

**ADHD Symptom Change for OTHER-EXP and OTHER-NAIVE**

An individual was classified as showing reliable symptom improvement if the difference between their CBCL ADHD Problems scores at Y2 and baseline was T<=-11. By this criterion, for those in the STIM-NAIVE group, 6 OTHER-EXP improved reliably and 18 did not, while 22 OTHER-NAIVE improved reliably and 52 did not. The 2X2 chi-squared test revealed no association between other exposure and reliable symptom improvement (𝜒^2^(1)=0.3, p=0.85).

**Change in rs-FC and Reliable Symptom Improvement for STIM-NAIVE**

As a confirmatory analysis, we tested for associations between ADHD symptom improvement based on the reliable change index and change in rs-FC for the three connections passing Step 2 (left caudate and frontoparietal network, left putamen and frontoparietal network, and right putamen and visual network) in STIM-NAIVE. We ran a Bayesian hierarchical logistic regression model identical to the one in Step 3 reported in the main text, predicting reliable symptom improvement. Results for these models did not reveal any evidence for associations between reliable symptom improvement and any connection: left caudate and frontoparietal network (Est.=2.27, sd=3.07, 95% QI=[-3.77,8.43], ESS=33991, Rhat=1.00), left putamen and frontoparietal network (Est.=0.21, sd=1.46, 95% QI=[-2.63,3.13], ESS=32880, Rhat=1.00), or right putamen and visual network (Est.=-1.44, sd=1.77, 95% QI=[-4.97,1.99], ESS=33841, Rhat=1.00). This was expected as the rs-FC connections included in Step 3 were selected for showing time-related change in STIM-EXP and not in STIM-NAIVE.
